# Prevalence of anti-NMDA receptor antibodies among adult Chinese patients presenting with first-episode psychosis in Hong Kong

**DOI:** 10.1101/2025.10.09.25337289

**Authors:** Steven WH Chau, Charis KW Lam, Matthew PM Yu, Joseph CH Choi, Howan HW Leung

## Abstract

**Background:** Studies on the presence of anti-NMDA receptor antibodies were done primarily on the European population. In addition, most of the existing studies did not collect samples during the acute phase of the psychosis. The prevalence of anti-NMDA receptor antibodies among the Asian population presenting with first-onset psychosis in an acute medical setting is under-investigated.

**Method:** We retrospectively recruited patients assessed by the consultation-liaison psychiatry team at an acute teaching hospital in Hong Kong, for first-onset psychosis who were unmedicated at the accident and emergency department or the acute medical wards from January 2015 to March 2025. Patients aged < 18 or > 65 were excluded. The presence of anti-NMDA receptor antibodies was tested by cell-based indirect immunofluorescence assay (EUROIMMUN) on serum and/or cerebrospinal fluid samples.

**Results:** We consecutively recruited 389 eligible patients. Among this population, 109 were tested for anti-NMDA receptor antibodies (Mean age = 36.0 years; 29.7% female; 97% ethnic Chinese, 82.6% diagnosed with primary psychotic disorders with no medical explanation). Cerebrospinal fluid (CSF) samples were taken from 60 of them. Among the tested patients, 2 (1.8%) were found to be positive for anti-NMDA autoantibodies (age 20s-30s; 1 female). The antibodies were detectable in both blood and CSF samples. Both of the patients exhibited the classic features of autoimmune encephalitis with marked CSF pleocytosis.

**Conclusion:** The prevalence of anti-NMDA receptor autoantibodies among our population presenting with first-episode psychosis is low, especially among patients with primary psychotic disorders. This prevalence is lower than that reported by studies mainly done among European-predominant populations. The prevalence of many known autoimmune diseases is known to vary among ethnic groups, and further studies are needed to explore the differences in anti-NMDA receptor positivity among these groups.

## Background

Anti-NMDA receptor antibodies are the most common type of anti-neuronal antibodies. Since its description in 2007 ^1^, the phenomenon of anti-NMDA receptor autoimmune encephalitis has been well described. The fact that this condition can present as acute psychosis means that this is an uncommon but important differential diagnosis for patients presenting with first-episode psychosis. Recent research has expanded the enquiry into the presence of antineuronal autoantibodies in patients presenting with psychosis but without encephalitis. Studies from the past decade report the prevalence of anti-NMDA receptor antibodies among patients with primary (or so-called ‘functional’) psychotic disorders to be around 1-4%, with considerable variability among the studies^2–4^. The role of anti-NMDA receptor antibodies and other antineuronal autoantibodies in the pathophysiology of psychiatric disorders in these patients remains debated.

However, the studies cited above were primarily conducted on the European population, and data from Asian populations are underrepresented. The epidemiology of autoimmune phenomena is known to be varied among ethnic groups. For example, multiple sclerosis is more common in the White population compared to the Asian population, while the reverse is true for systemic lupus erythematosus^5,6^. In addition, most of the existing studies did not collect samples during the acute phase of psychosis. For example, the only available study examining the prevalence of anti-NMDA receptor antibodies among Chinese with first-episode psychosis was done in the setting of a specialist outpatient for early psychosis in Hong Kong^7^, which would have excluded patients with a diagnosis of autoimmune encephalitis.

The primary objective of the current study is to investigate the prevalence of anti-NMDA antibodies among Chinese adult patients presenting with unmedicated first-onset psychosis in an acute medical setting. The secondary objective is to explore the presentation of patients with psychosis and anti-NMDA receptor antibodies.

## Method

This is a retrospective cross-sectional study. We reviewed all the cases being assessed by the consultation liaison psychiatry team at the Accident and Emergency Department or inpatient departments of is a major tertiary hospital of Hong Kong serving a catchment area of a population of 700 thousand people, for first-onset psychosis from January 2015 to March 2025. The inclusion criteria are: 1. Age 18-65 years old at the time of presentation; 2. Presenting with first-onset psychosis, regardless of etiologies; 3. Received test(s) for the presence of anti-NMDA receptor antibodies in serum or cerebrospinal fluid sample. The presence of anti-NMDA receptor antibodies was tested by cell-based indirect immunofluorescence assay (EUROIMMUN) in a certified laboratory of another teaching hospital. The study was approved by the Joint CUHK-NTEC CREC (ref no.: 2024.606).

## Result

Among the 426 patients assessed for suspected first-onset psychosis over the recruitment period, 389 were confirmed to have first-onset psychotic symptoms. 109 of them were tested for anti-NMDA receptor antibodies (Mean age = 36.0 years; 71.6% female; 96% ethnic Chinese). The selection of patients for antibody testing was based on clinical suspicion; therefore, the patients tested had significantly different demographic and clinical characteristics, such as age, acuteness, presence of fever, and catatonia. (See Table 1). Cerebrospinal fluid (CSF) samples were tested in 60 of them. The comparison of the demographic and clinical characteristics of the patients of first-onset psychosis with and without anti-NMDA receptor autoantibodies testing is shown in Table 1. Among the tested patients, 35 (32%) were diagnosed with non-specific psychosis, 21 (19%) were diagnosed with schizophrenia, 15 (14%) were diagnosed with acute and transient psychotic disorder, 10 (9%) were diagnosed with mood disorder with psychotic symptoms and 19 (17%) patients were diagnosed with psychosis secondary to substance use or other medical conditions. Two of the patients (1.8%) were found to be positive for anti-NMDA receptor antibodies (ages 20s to 30s; 1 female, both are Chinese). The antibodies were detectable from both blood and CSF samples in both patients, and they both exhibited the classic clinical features of autoimmune encephalitis with marked CSF pleocytosis: The first patient presented with abrupt-onset insomnia, cognitive impairment, and auditory and visual hallucinations within a week. The patient subsequently developed orofacial dyskinesia and seizures in the following two weeks. While the MRI of the brain with contrast did not reveal encephalitic changes, the CSF analysis showed marked pleocytosis without evidence of infection. The second patient presented with an acute onset of cognitive impairment, agitation, and insomnia for two weeks, followed by the onset of auditory hallucination two days before hospitalization. CSF analysis of the patient showed marked pleocytosis without evidence of infection. The MRI brain scan showed an abnormal T2W signal over the left temporal lobe, suggestive of encephalitis, which resolved after treatment. In both patients, extensive work-ups, including full-body PET-CT scans, did not reveal any malignancy. Both patients’ neuropsychiatric symptoms responded to aggressive immunotherapy.

**Table 1.** Comparison of demographical and clinical characteristics between patients with first-episode psychosis tested for anti-NMDA receptor antibodies and those untested.

|  | **FEPs without anti NMDA-R ab tested (n= 280)** | **FEPs with anti NMDA-R ab tested (n= 109)** | **P-value** |
| --- | --- | --- | --- |
| Mean age(SD), years | 41.6(13.5) | 36.0(12.8) | <0.001<sup>a</sup> |
| Female, % | 67.9% | 71.6% | 0.48<sup>b</sup> |
| Non-ethnic Chinese, % | 2.9% | 3.7% | 0.76<sup>c</sup> |
| Onset < 1 month, % | 27.1% | 56.8% | <0.001<sup>b</sup> |
| Presence of fever ($\geq 37.5^{\circ}\text{C}$), % | 20.0% | 43.1% | <0.001<sup>b</sup> |
| Catatonia at presentation, % | 1.8% | 8.3% | 0.004<sup>c</sup> |

a: t-test was used; b: Chi-square test was used; c: Fisher's exact test was used.

## Discussion

To the best of our knowledge, this is the first study investigating the prevalence of anti-NMDA receptor antibodies among patients presenting with first-episode psychosis in an acute medical setting in a predominantly Chinese population.

Our results suggest that the prevalence of anti-NMDA receptor antibodies among patients with first-onset psychosis is low among our cohort. Particularly, we do not find any antibody-positive case among patients suffering from primary psychotic disorders. A previous study on the prevalence of anti-NMDA receptor antibodies among patients with psychotic disorders in an outpatient setting in Hong Kong Chinese also yielded a low prevalence of 1.5%^7^. The prevalence reported in the literature, predominantly based on the Western population, has been highly variable. This variability can be partly attributed to diverse sampling contexts among the studies. The different kinds of assay used by the study could also explain part of the variability, as live cell-based assay is shown to be more sensitive than fixed cell assay, especially for serum test^8^. However, whether ethnic disparity plays a part in the variability is under-investigated. A recent multi-center study from Europe, which reported the prevalence of anti-NMDA receptor antibodies in their cohort by ethnicity, showed marked variations in prevalence among the ethnic groups^9^. Thus, we hypothesize that there is a potential ethnic disparity in the prevalence of anti-NMDA receptor antibodies among patients with psychosis, and further studies are needed to test this hypothesis. The limitations of the current study include the small sample size and its single-center design. While the study samples were consecutively recruited, the selection of patients undergoing the test for anti-NMDA receptor antibodies is biased by clinical suspicion. However, such bias is more likely to result in an overestimation of the prevalence. The age-related inclusion criteria, which are a restriction due to the scope of our team’s service, mean that potentially some cases of young-onset psychosis or encephalitis are missed from the study.

## Data Availability

All data produced in the present study are available upon reasonable request to the authors

